# Corticosteroids are associated with increased survival in elderly presenting severe SARS-Cov2 infection

**DOI:** 10.1101/2020.11.10.20226886

**Authors:** Gallay Laure, Tran Viet-Thi, Perrodeau Elodie, Vignier Nicolas, Mahevas Matthieu, Bisio Francesca, Forestier Emmanuel, Lescure Francois-Xavier, on behalf of the COCO-OLD Study group

**Author notes:** **Corresponding author:** Lescure FX, MD PhD, Service de maladies infectieuses et tropicales, Hôpital Bichat, Assistance Publique Hôpitaux de Paris, 75018 Paris, France. COCO-OLD Study Group (file attached).

## Abstract

**Objective:** To assess the effectiveness of corticosteroids among elderly patients with COVID-19 pneumonia requiring oxygen.

**Design:** Comparative observational study based on routine care data. Baseline characteristics of patients were balanced using propensity-score inverse probability of treatment weighting.

**Setting:** Geriatric and infectious diseases wards from 36 hospitals in France and Luxembourg.

**Participants:** Adults > 80 years old PCR confirmed SARS-CoV-2 infection or typical CT-scan images, requiring oxygen ≥ 3L/min and with an inflammatory syndrome (C-reactive protein ≥ 40mg/L).

**Measurements:** The primary outcome was overall survival at day 14. The secondary outcome was the proportion of patients discharged from hospital to home/rehabilitation on day 14. Adverse events were abstracted from electronic health records.

**Results:** Among the 267 patients included in the analysis, 96 were assigned to the treatment group. Median age was 86, interquartile range 83 to 90 and 95% had a SARS-CoV-2 PCR-confirmed diagnosis. Use of corticosteroids was significantly associated with an increased survival (weighted hazard ratio [wHR] 0.66, 95% CI 0.44 to 0.97). There was no significant difference between the treatment and control groups regarding the proportion of patients discharged to home/rehabilitation at day 14 (wRR 1.11, 95% CI 0.68 to 1.81). Twenty-two (16.7%) patients receiving corticosteroids developed adverse events while only 11 (6.4%) from the control group did.

**Conclusions:** Corticosteroids were associated with a significant increase the day-14 overall survival of patients over 80 years old hospitalized for severe COVID-19.

**Impact statement:** We certify that this work is novel. As of today, studies on the efficacy of corticosteroids did not specifically target elderly patients.

Among older patients aged over 80 years old, the RECOVERY trial found no difference in the survival of patients treated or not with dexamethasone. However, the heterogeneity in the severity of infection within the latter subgroup limited the drawing of strong conclusions

## Background

Older adults are at greatest risk of severe disease and death due to the coronavirus disease 2019 (COVID-19). Globally, people older than 65 years comprise 9% of the population, yet account for 30% to 40% of cases and more than 80% of COVID-19-related deaths ^1^. Although the WHO recently recommended treatment with systemic corticosteroids for all adult patients with severe and critical COVID-19 ^2^, the evidence supporting this recommendation for patients over 80 years old are weak. Indeed, among these patients, the RECOVERY trial found no difference in the survival of patients treated or not with dexamethasone ^3^. However, the heterogeneity in the severity of infection within the latter subgroup limits the drawing of strong conclusions ^4^. This situation has raised concerns and, in some countries, such as France, recommendations do not support the systematic use corticosteroids for patients ≥ 70 years old ^5^.

Given the high lethality of COVID-19 among older patients and the shortage of ICU beds, corticosteroids have been frequently administered at the beginning of the epidemic in France to patients aged 80 years or more. We aimed at using these data to retrospectively emulate a target trial, with the objective of assessing the effectiveness of corticosteroids as treatment for severe SARS-CoV-2 infection in elderly patients.

## Methods

### Participants and settings

Data collected from 36 French non-ICU medical wards were used to emulate a target trial assessing the effectiveness of corticosteroids in elderly patients with COVID-19 using methods previously described in ^6^. All patients hospitalized between March 1^st^ and April 30^th^ 2020 who were aged 80 years or more, who suffered from a SARS-CoV-2 infection confirmed by PCR or by typical clinical and CT-scan findings, who required oxygen therapy ≥3 L/min, and who had C-reactive protein levels ≥40 mg/L, were included in the study.

### Treatment strategies

We compared two treatment strategies: receiving at least one dose of corticosteroids ≥0.4 mg/kg/day eq. prednisone (treatment group), or receiving the standard of care (control group). The cut-off value of 0.4mg/kg/day was chosen to account for dose rounding by physicians who had the intent to treat patients with corticosteroids at 0.5 mg/kg/day eq. prednisone. To emulate a pragmatic trial, patients in the treatment group could start corticosteroids within a “grace period” of 72h after baseline.

### Follow-up

The start of follow-up (baseline or time zero) for each individual was the time all eligibility criteria (oxygen therapy ≥ 3L/min and inflammatory syndrome with a CRP level ≥ 40mg/L) were met. All patients were followed up from baseline until whichever of the following events occurred first: (1) death, (2) loss to follow-up, or (3) end of follow-up, which occurred at least 14 days after baseline.

### Outcomes

The primary outcome was overall survival at day 14. The secondary outcome was the proportion of patients discharged from hospital to home/rehabilitation on day 14. All adverse events were abstracted from electronic health records in free text and independently recoded by one physician (VTT).

### Statistical analyses

Our causal contrast of interest was the per-protocol effect and we compared characteristics of participants who received corticosteroids within 72 hours from baseline to those who did not receive the drug. We thus excluded from the analysis the patients who started corticosteroids before baseline, the patients who received corticosteroids >72h after baseline, and the patients who received corticosteroids at a dose lower than 0.4mg/kg per body weight.

Propensity-score methods were used to account for differences between the two groups at baseline. The propensity score represents the probability for patients to receive corticosteroids given their baseline demographic and clinical covariates (Details on the propensity score development are available in the **Supplementary materials**). Estimates of the average treatment effect were calculated by inverse probability of treatment weighting (IPTW) using cox proportional hazards models for hazard ratios (wHR) and IPTW estimates of the relative risk (wRR) for binary outcomes.

To account for immortal time bias, all patients in the control group who died during the grace period were randomly assigned to one of the two groups, given that their observational data were compatible with both groups at the time of the event ^7^.

Missing baseline variables were handled by multiple imputation by chained equations. Methods are fully described in the Supplementary materials. The study was approved by the IRB of the Henri Mondor Hospital (2020_060).

## Results

### Participants

A total of 267 patients included, their median age was 86 years (interquartile range [IQR], 83 to 90), 49.8% were men, 95% had a SARS-CoV-2 PCR-confirmed diagnosis, and 96 were assigned to the treatment group. Both comorbidities and clinical severity at baseline were similar between the two groups. However, patients with low autonomy at baseline, measured using the GIR score, were less often prescribed corticosteroids (14.6% and 20.6% of patients had a GIR score of 1 or 2 in the treatment and control group, respectively).

The median time from baseline to the symptoms onset was 7 days (IQR, 4 to 10). Among the 96 patients assigned to the treatment group, 51 (53.7%) received methylprednisolone, 22 (23.2%) received prednisone, 15 (15.8%) received dexamethasone, 4 (4.2%) received prednisolone, and 3 (3.2%) received hydrocortisone (**Table**).

### Follow-up and outcomes

In total, 41 (42.7%) and 86 (50.2%) patients died before day 14 in the treatment and control groups, respectively (HR 0.76, 95% CI 0.52 to 1.09). After balancing the baseline covariates by IPTW, the survival was significantly higher in for patients from the treatment group compared to those from the control group, wHR 0.66, 0.44 to 0.97 (**Figure**). There was no significant difference between the treatment and control groups regarding the proportion of patients discharged to home/rehabilitation at day 14 (wRR 1.11, 95% CI 0.68 to 1.81).

Twenty-two (16.7%) patients receiving corticosteroids developed adverse events while only 11 (6.4%) from the control group did: hyperglycemia [6.1% vs. 0.6%], heart failure [2.3% vs. 0.6%], confusion [3.0% vs. 1.2%], infection [1.5% vs. 0%].

## Discussion

In the present emulated trial using real-life data, corticosteroids significantly improved the 14- days-survival of patients aged 80 years or more and hospitalized in a non-ICU department for severe COVID-19.

Recommendations to treat older patients with corticosteroids have been based on results from studies performed in ICU ^8^ and on results from the RECOVERY trial, which showed an overall effect of dexamethasone, but a non-significant difference in the population of patients aged 80 years or more. The discrepancy between the results from the RECOVERY trial and ours may be explained by the heterogeneity in treatment effect. Indeed, for patients from the RECOVRY trial who were not under oxygen at the time of randomization (35% of patients older than 80 years) corticosteroids were not associated with benefit. For those with oxygen at the time of randomisation, corticosteroids showed a significant reduction in mortality. In our study, the population was more homogenous and all patients required oxygen and had a severe - but not critical - disease according to WHO criteria. Our findings fill the evidence gap for this specific and vulnerable population and support the WHO guidelines to recommend corticosteroids for patients with severe and critical COVID 19 (defined by any of oxygen saturation < 90% on room air, respiratory rate > 30 breaths per minute in adults, and/or signs of severe respiratory distress).

Besides survival, corticosteroids might also shorten the duration of symptoms and/or hospitalization, and therefore reduce the geriatric complications resulting from SARS-CoV2 infection; such effect was not observed in the present study, although the end point at day 14 might have been too early to objectivize this. Decreasing functional decline, nutrition, and cognitive impairment is a major issue during the management of COVID-19 and other infectious diseases in the elderly population ^9^. Further studies should be performed to explore this crucial point.

Regarding tolerance, patients treated with corticosteroids experienced more adverse events than those treated with the standard of care. Nevertheless, those effects remained rare and there was no major toxicity, especially cardio- or neurotoxicity. WHO experts stated that potential side effects of corticosteroids are well balanced by their positive effect on mortality. Moreover, clinicians are used to manage corticosteroids and their side effects. Our results support this statement.

The present study suffered several limitations. Despite the use of robust methods and statistical techniques to draw causal inferences, the study remains observational and potential unmeasured confounders may bias the results ^10^. Also, the prescriptions of corticosteroids were heterogeneous in terms of drugs, time of start, dose, and duration. Finally, the follow-up was limited at 14 days. Yet, deaths occurred mainly before day 10, and patients particularly frail were managed frequently in a rescue context without practicing intensive care.

Altogether, our results support the WHO guidelines, and expand them to patients over 80 years old without contra-indication. They send a good signal for elderly, including those living in long terms care facilities or other institutions where corticosteroids could be prescribed according to an oxygen criterion, without systematic transfer to the hospital. Strengthening the therapeutic arsenal for the care of elderly with COVID-19 is critical as these patients may not fully benefit from vaccination because of the immune-senescence associated with advanced age and their exclusion from vaccine trials ^4^.

## Supporting information

Study group

## Data Availability

Data are available upon reasonable request

## Author Contributions

Concept and design: Lescure FX, Gallay L, Tran VT.

Acquisition, analysis, or interpretation of data: All authors from the COCO_OLD Study Group.

Drafting of the manuscript: Tran VT, Lescure FX, Forestier E, Gallay L, Bisio F, Vignier N., Mahevas M.

Critical revision of the manuscript for important intellectual content: All authors. Statistical analysis: Tran VT, Perrodeau E.

Administrative, technical, or material support: Lescure FX, Gallay L, Tran VT, Forestier E. Supervision: Lescure FX.

Dr Tran and Ms. Perrodeau had full access to all the data of the study and take responsibility for the integrity of the data and the accuracy of the data analysis.

---

**Table:**
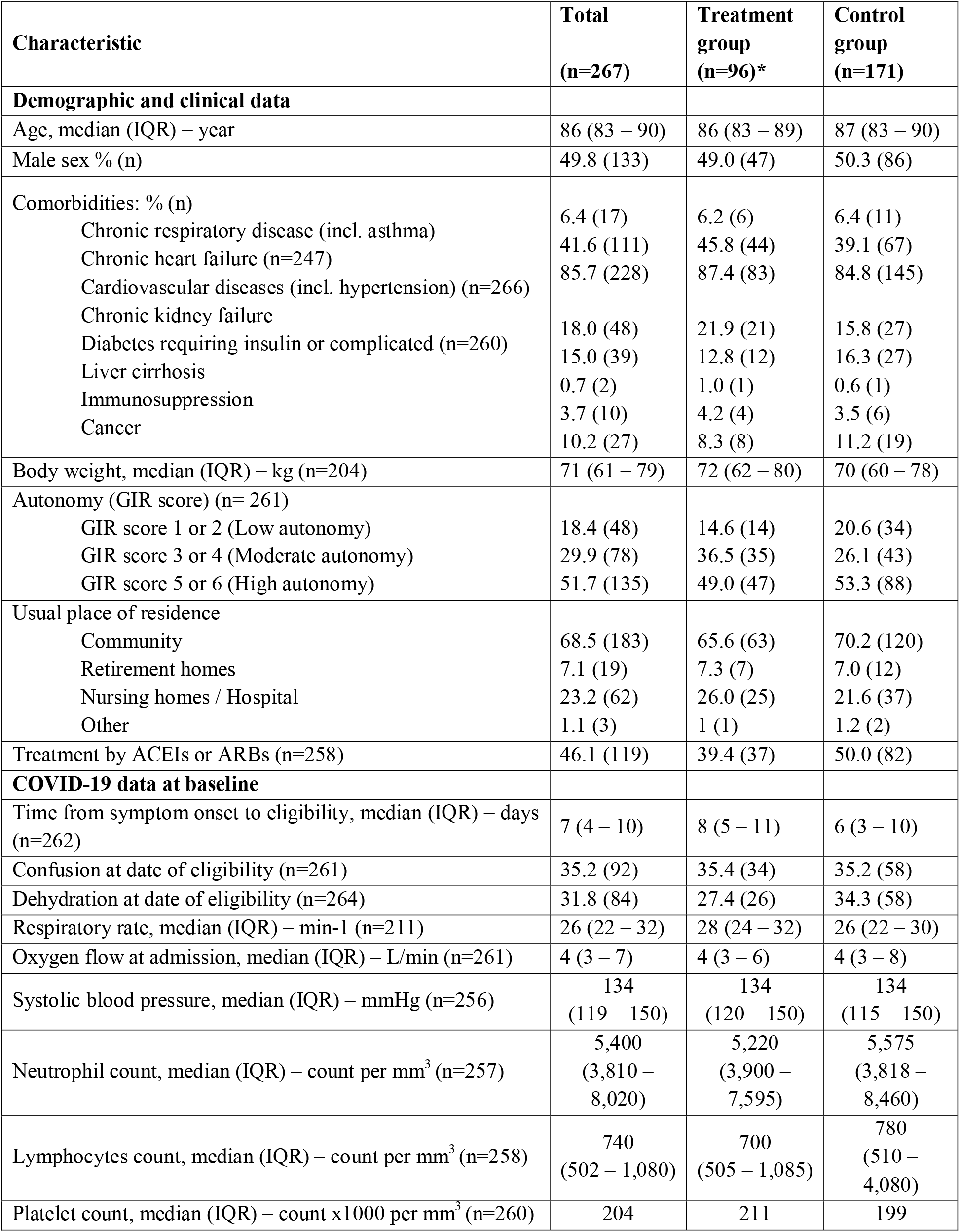
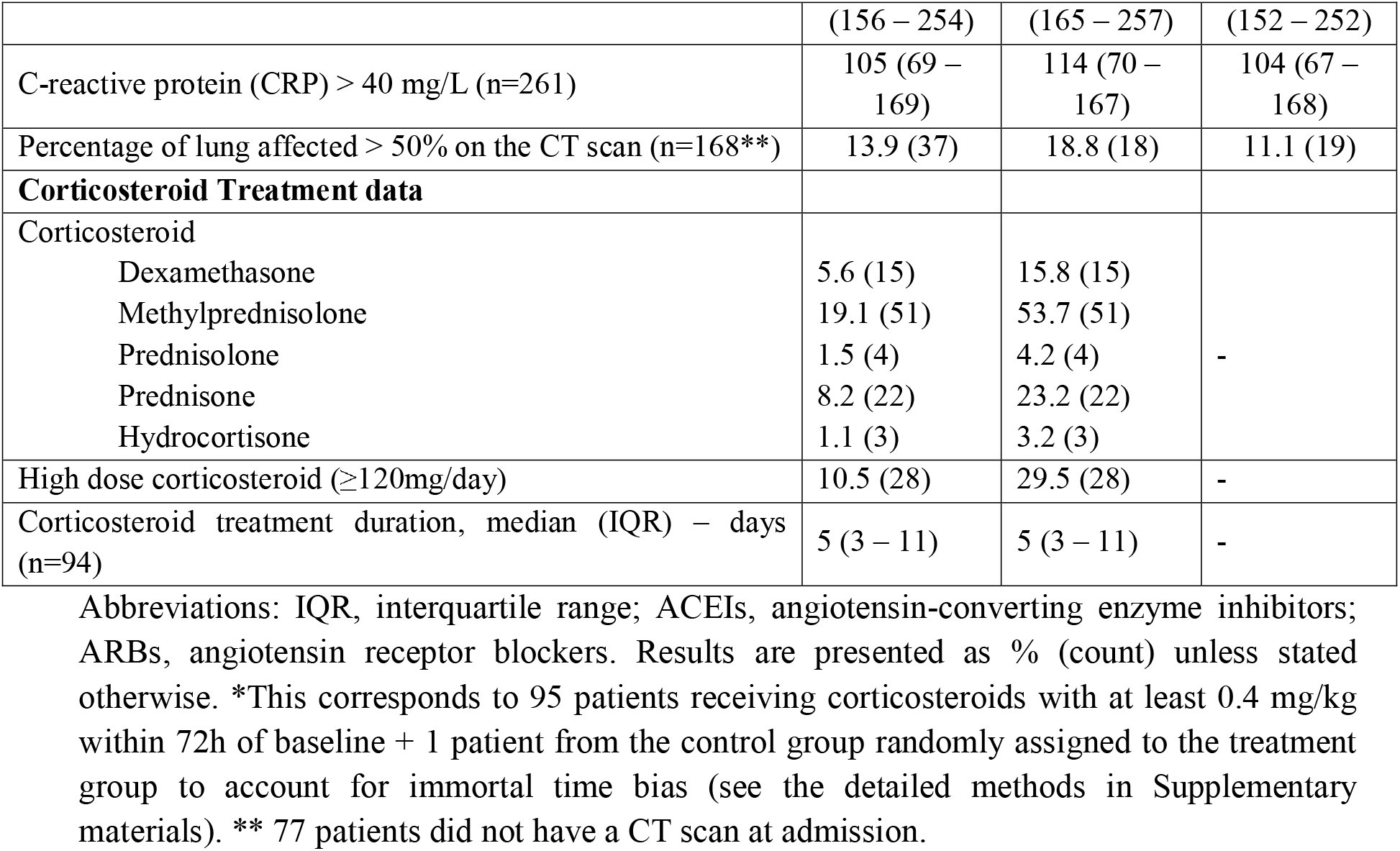
patients’ baseline characteristics (n=267). Number in brackets in the first column correspond to the quantity of data available before imputation of missing baseline data by multiple imputations by chained equations.

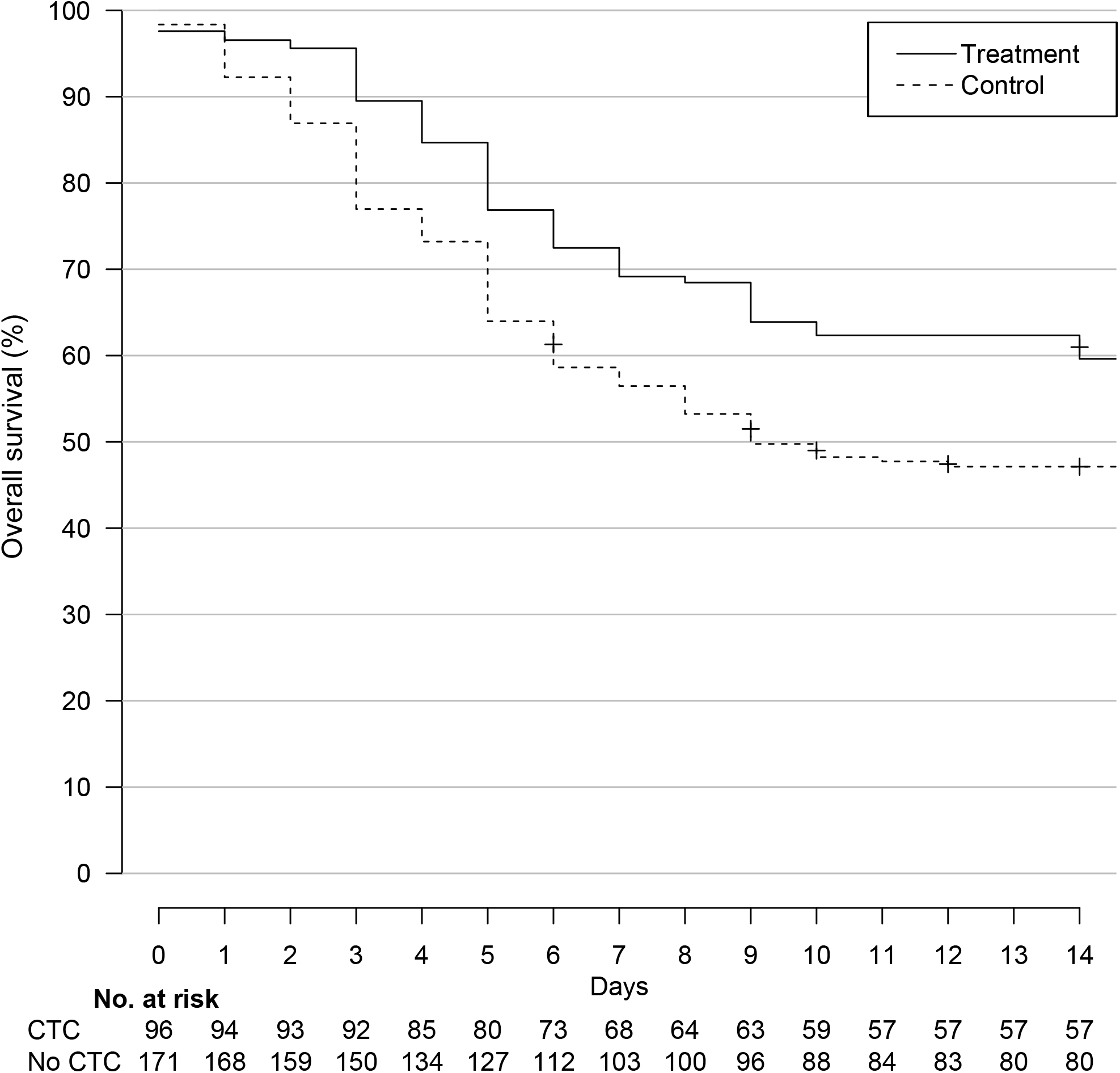

