## Supplementary material for "Corticosteroids are associated with increased survival in elderly presenting severe SARS-Cov2 infection": Study group

### COCO-OLD (Collaborative cOhort COrticoteroids for OLD patients with COvid-19) study Group

### **Principal investigator**

François-Xavier Lescure

### Methodology and statistics

Centre d’Epidémiologie Clinique, Hôpital Hôtel-Dieu, Assistance Publique-Hôpitaux de Paris AP-HP / Université de Paris, Centre de Recherche Epidémiologie et Statistiques (CRESS UMR 1153) : Viet-Thi Tran, Elodie Perrodeau

### Centres (alphabetically)

**Ales, centre hospitalier, Service de médecine interne** : Thibaut Fraisse

**Angers, centre hospitalo-universitaire, Service de Maladies Infectieuses et Tropicales** : Diane Sanderink

**Blois, centre hospitalier, Service de médecine interne et Maladies infectieuses** : Bertrand Lioger

**Cayenne, centre hospitalier, Service Maladies infectieuses et tropicales :** Camille Boutrou

**Chambéry, Centre hospitalier métropole Savoie, Service Maladies infectieuses et tropicales :** Anne - Laure Destrem

**Chateaubriand, centre hospitalier, Service médecine polyvalente et gériatrie aigue :** Pascal Gicquel

**Colmar, hôpital Louis Pasteur, Service de maladies infectieuses et tropicales :** Martin Martinot

**Fort de France, centre hospitalo-universitaire Martinique, Service des maladies infectieuses et tropicales :** Jérémie Pasquier

**Luxembourg (Luxembourg), centre hospitalier, service de** **réanimation polyvalente**: Jean Reuter

**Lyon Hospices civils de Lyon, hôpital Edouard Herriot, médecine interne** : Helene Desmurs-Clavel

**Lyon, Hospices civils de Lyon, hôpital de la Croix Rousse, Maladies Infectieuses et Tropicales** : Nicolas Benech

**Marseille, hôpital St Joseph, Service de Médecine Interne** : Boris Bienvenu

**Melun, Centre hospitalier,** **Service Maladies infectieuses et tropicales** : Nicolas Vignier

**Montreuil, centre hospitalier intercommunal André Grégoire, Service de médecine interne et Maladies infectieuses** : Guillemette Frémont

**Nancy, centre hospitalo-universitaire, hôpital Brabois, Maladies Infectieuses et Tropicales** : François Goehringer

**Nantes, centre hospitalo-universitaire, service de Médecine Aiguë Gériatrique** : Guillaume Chapelet

**Nantes, Hôpital privé du confluent de Nantes, Service de Médecine Interne et Maladies Infectieuses :** Olivier Grossi

**Nîmes, centre hospitalo-universitaire Carémeau,** **Service des maladies infectieuses et tropicales** : Didier Laureillard

**Paris, Assistance Publique Hôpitaux de Paris (AP-HP), Hôpital Bichat :** Cyrille Gourjault, Alexandre Lahens, François-Xavier Lescure

**Paris, Assistance Publique Hôpitaux de Paris (AP-HP), Hôpital Cochin, Service de Médecine interne** : Célia Azoulay

**Paris, Assistance Publique Hôpitaux de Paris (AP-HP), Hôpital Cochin, Service de pneumologie** : Nicolas Carlier

**Paris, Assistance Publique Hôpitaux de Paris (AP-HP), Hôpital Pitié Salpêtrière, Service des Maladies Infectieuses et Tropicales** : Gianpiero Tebano

**Paris, Assistance Publique Hôpitaux de Paris (AP-HP), Hôpital St Antoine** : Jérôme Pacanowski

**Paris, Assistance Publique Hôpitaux de Paris (AP-HP), Hôpital Jean Verdier** : Simone Tunesi

**Paris, Assistance Publique Hôpitaux de Paris (AP-HP), Hôpital Tenon :** Nadège Lemaire

**Pontivy, centre hospitalier centre Bretagne,** **Service de Médecine Polyvalente, Maladies Infectieuses, Dermatologie** : Laurent Bellec

**Reims, centre hospitalo-universitaire, service de Médecine interne, Immunologie clinique et maladies infectieuses** : Firouze Bani-Sadr

**Roubaix,** **centre hospitalier, Service de Médecine interne** : Marie Pichenot

**Rouen, centre hospitalo-universitaire, Service des Maladies Infectieuses et Tropicales** : Kevin Alexandre

**Saint Denis (réunion),** **centre hospitalo-universitaire, service de pneumoligie** : Laurie Masse

**Tourcoing, centre hospitalier, hôpital Guy Chatiliez, Service Universitaire des Maladies Infectieuses et du Voyageur** : Olivier Robineau

**Tours, centre hospitalo-universitaire, Pôle Médecine** : Camille Thorey, Sophie Deriaz

**Valence, centre hospitalier, Service de maladie infectieuse** : Julien Saison

**Vannes, Centre Hospitalier Bretagne Atlantique,** **Service de médecine interne, maladies infectieuses et hématologie :** Marie Gousseff

**Versaille,** **centre hospitalier, Service médecine polyvalente et gériatrie aigue** : Laura Goehrs

**Villejuif, Institut Gustave Roussy, Département Interdisciplinaire d’Organisation des Parcours Patients**: Fanny Pommeret

**Vierzon, centre hospitalier, Service de maladie infectieuse** : Francesca Bisio
