## Supplemental files for "Corticosteroids are associated with increased survival in elderly presenting severe SARS-Cov2 infection"

### Supplementary material : Propensity score development

We used an inverse probability of treatment weighting (IPTW) approach based on patients’ propensity scores (i.e. patients’ predicted probability of receiving a certain CTC given their baseline covariates) to balance the differences in baseline variables between treatment groups.

A non-parsimonious multivariable logistic regression model was constructed to estimate each patient’s propensity score. Variables of the propensity score (PS) model were planned and prespecified before any outcome analyses, and included:

- Age
- Gender
- Autonomy before hospitalization (measured using the GIR score)
- Usual living environment (home, retirement home, etc.)
- Presence of a chronic respiratory insufficiency or a chronic respiratory pathology likely to decompensate during a viral infection
- Heart failure; chronic kidney disease
- Liver cirrhosis
- Personal history of cardiovascular disease [hypertension, stroke, coronary artery disease, or cardiac surgery]
- Insulin-dependent diabetes mellitus, or diabetic microangiopathy or macroangiopathy
- Immunosuppression (because of immunosuppressive drugs, including anticancer chemotherapy)
- Uncontrolled HIV infection or HIV infection with CD4 cell counts < 200/µL; or a haematological malignancy
- Body weight
- Treatment by angiotensin-converting enzyme inhibitors (ACEIs) or angiotensin receptor blockers (ARBs)
- Time since symptom onset
- Percentage of lung affected on the CT scan
- Presence of confusion
- Respiratory frequency
- Oxygen flow at inclusion
- Systolic blood pressure
- CRP

All variables included in the propensity score model reflected the knowledge available at baseline.

Standardised differences were examined to assess balance, with a threshold of 10% designated to indicate clinically meaningful imbalance.

We present here the propensity scores for the treatment and standard of care groups (n=267) before (panel A) and after IPTW (panel B).

A

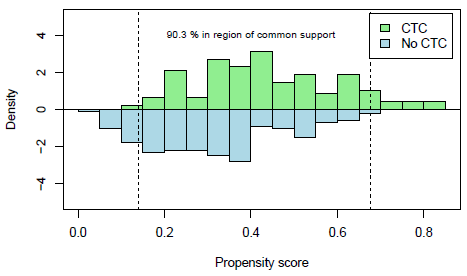

**B**

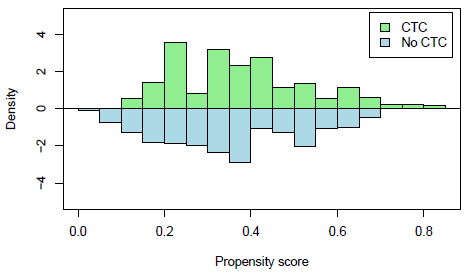

CTC: corticosteroid treatment; IPTW: inverse probability of treatment weighting.

We present below the standardised differences of variables used to generate the propensity score before and after IPTW (n=267).

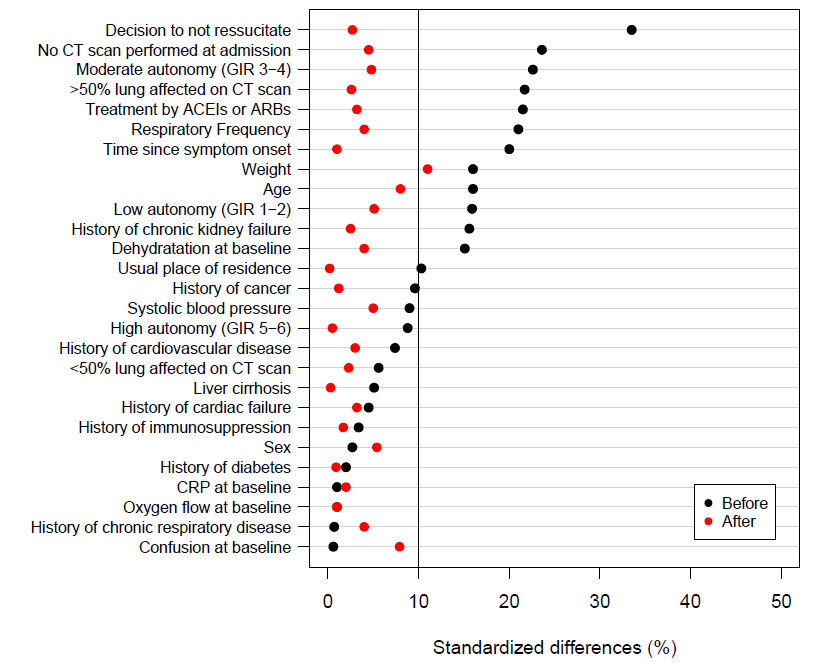
